# Advanced Restriction imaging and reconstruction Technology for Prostate MRI (ART-Pro): Study protocol for a multicenter, multinational trial evaluating biparametric MRI and advanced, quantitative diffusion MRI for detection of prostate cancer

**DOI:** 10.1101/2024.08.29.24311575

**Authors:** Madison T Baxter, Christopher C Conlin, Aditya Bagrodia, Tristan Barrett, Hauke Bartsch, Anja Brau, Matthew Cooperberg, Anders M Dale, Arnaud Guidon, Michael E Hahn, Mukesh G Harisinghani, Juan F Javier-DesLoges, Sophia Kamran (Capuano), Christopher J Kane, Joshua M Kuperman, Daniel JA Margolis, Paul M Murphy, Nabih Nakrour, Michael A Ohliger, Rebecca Rakow-Penner, Ahmed Shabaik, Jeffry P Simko, Clare M Tempany, Natasha Wehrli, Sean A Woolen, Jingjing Zou, Tyler M Seibert

## Abstract

**Background:** Multiparametric MRI (mpMRI) is strongly recommended by current clinical guidelines for improved detection of clinically significant prostate cancer (csPCa). However, major limitations of mpMRI are the need for intravenous (IV) contrast and dependence on reader expertise. Efforts to address these issues include use of biparametric MRI (bpMRI) and advanced, quantitative MRI techniques. One such advanced technique is the Restriction Spectrum Imaging restriction score (RSIrs), an imaging biomarker that has been shown to improve quantitative accuracy of patient-level csPCa detection.

**Purpose:** To evaluate whether IV contrast can be avoided in the setting of standardized, state-of-the-art image acquisition, with or without addition of RSIrs, and to evaluate characteristics of RSIrs as a stand-alone, quantitative biomarker.

**Design, setting, and participants:** ART-Pro is a multisite, multinational trial that will be conducted in two stages, evaluating bpMRI, mpMRI, and RSIrs on accuracy of expert (ART-Pro-1) and non-expert (ART-Pro-2) radiologists’ detection of csPCa. Additionally, RSIrs will be evaluated as a stand-alone, quantitative, objective biomarker (ART-Pro-1). This study will include a total of 500 patients referred for a multiparametric prostate MRI with a clinical suspicion of prostate cancer at any of the five participating sites (100 patients per site).

**Intervention:** In ART-Pro-1, patients receive standard of care mpMRI, with addition of the RSI sequence, and subsets of the patients’ images are read separately by two expert radiologists, one of whom is the standard of care radiologist (Reader 1). Three research reports are generated using: bpMRI only (Reader 1), mpMRI (Reader 1), and bpMRI + RSIrs (Reader 2). The clinical report is submitted by Reader 1. Patients’ future prostate cancer management will be recorded and used to evaluate the performance of the MRI techniques being tested.

In ART-Pro-2, the dataset created in ART-Pro-1 will be retrospectively reviewed by radiologists of varying experience level (novice, basic, and expert). Radiologists will be assigned to read cases and record research reports while viewing subsets of either mpMRI only or RSIrs + mpMRI. Patient cases will be read by two readers from each experience level (6 reads total), and findings will be evaluated against the expertly created dataset from ART-Pro-1.

**Outcome measurements and statistical analysis:** The primary endpoint is to evaluate if bpMRI is non-inferior to mpMRI among expert radiologists (ART-Pro-1) and non-expert radiologists (ART-Pro-2) for detection of grade group (GG) ≥2 csPCa. We will conduct one-sided non-inferiority tests of correlated proportions (ART-Pro-1) and use McNemar’s test and AUC to test the null hypothesis of non-inferiority (ART-Pro-1 and ART-Pro-2).

**Conclusions:** This trial is registered in the US National Library of Medicine Trial Registry (NCT number: NCT06579417) at ClinicalTrials.gov. Patient accrual at the first site (UC San Diego) began in December 2023. The expected trial timeline is three years to complete accrual with a six-month endpoint.

## 1. Introduction

Clinical guidelines strongly recommend multiparametric MRI (mpMRI) to improve detection of clinically significant prostate cancer (csPCa) and to avoid unnecessary biopsies^1–4^. With prostate cancer (PCa) diagnoses currently at 1.4 million per year in 2020 and expected to double by 2040—and considering that millions more men will be evaluated for possible cancer—there is a critical need to dramatically increase capacity for prostate MRI^5^. Already, access to prostate MRI is limited, creating a health disparity that often disproportionately affects those at highest risk of dying from PCa^6,7^. Major limitations to scaling up capacity for mpMRI prior to biopsy are dependence on reader expertise and the need for intravenous (IV) contrast.

Prostate mpMRI interpretation is dependent on reader expertise and inherently subjective. Despite guidelines to standardize image acquisition and reporting (PI-RADS)^8^, results vary widely between radiologists and imaging centers^9,10^. To achieve good results with mpMRI, radiologists must gain significant training and experience^11^, specifically for prostate MRI^12^, which often requires years and considerable resources. A rapid increase in the supply of expert prostate radiologists may not be feasible, and while AI very likely will help fill this void, development of such tools will require large, well annotated, standardized patient cohorts to develop and validate such AI-based tools. Changes in imaging technology (e.g., scanners, operating systems, and/or reconstruction methods) may also have unpredictable effects on deep-learning AI models. Imaging quality is also variable, a reflection on both the heterogeneity of MRI equipment and the experience of imaging center staff in designing and following prostate cancer acquisition protocols. Though PI-RADS does list some technical specifications, there is still a nearly unlimited range of permissible protocols for acquiring prostate MRI data, even on the same scanner^8^. Vendor, scanner model, and operating system versions add further opportunity for complexity. Additionally, many prostate MRI scans are not compliant with the minimal PI-RADS technical specifications. Two multisite studies evaluating adherence to individual PI-RADS acquisition parameters found that fewer than 20% of scans were compliant^13,14^. Further standardization of prostate MRI protocols beyond meeting the minimal PI-RADS specifications, for example, using PI-QUAL^15^, has potential to greatly improve consistency of images.

Biparametric MRI (bpMRI) is mpMRI without IV contrast. bpMRI avoids the issues associated with invasive contrast injection, including increased patient risk, limited accessibility, and increased time and cost. First, obtaining intravenous access requires a skilled technologist or nurse. Additionally, because administration of contrast presents risks to patients, skilled supervision is required; in the U.S., physicians are required to be present during administration, which may limit the ability to scan patients outside normal working hours or in remote or underserved areas. The dynamic contrast enhanced (DCE) MRI sequence also requires additional scan time, which reduces scanner availability. Thus, alleviating the need for IV contrast by use of bpMRI affords the advantages of improving patient comfort, reducing cost, saving time, and increasing capacity. There is evidence that bpMRI may be comparable to mpMRI for guiding biopsy decisions^16^, though results are mixed. First, when using bpMRI only, radiologists tend to find more false-positive lesions, leading to unnecessary biopsies^17^. Second, DCE helps non-expert radiologists detect suspicious regions they could otherwise miss with bpMRI, resulting in overall higher sensitivity compared to bpMRI alone^18^. Third, DCE also often serves as a backup when diffusion-weighted imaging (DWI) is of poor quality^17,19^, so to avoid the use of contrast, DWI needs to consistently be of high quality. Overall, bpMRI may potentially remove one barrier (IV contrast) to increased prostate MRI capacity while exacerbating another barrier (dependence on exam quality and radiologist expertise).

Prostate MRI quality may greatly benefit from new MR technological advances. Scanner hardware continues to improve, including addition of surface coils that can be placed on the patient to yield higher signal to noise. Modern scanners with higher field strength (3.0 tesla) do not require an endorectal coil to be inserted into the patient. Higher gradient performance yields better DWI, a critical part of bpMRI and mpMRI. Software advances, too, can make important contributions to image quality. Reconstruction techniques that leverage deep-learning artificial intelligence (AI) enable rapid acquisition of high-quality images^20^. Another approach to mitigate image quality issues is to standardize acquisition protocols for prostate MRI across centers, at least for a given vendor and software version.

Restriction Spectrum Imaging (RSI) is an advanced diffusion technique that can generate images with high specificity for csPCa^21–24^. RSI can be efficiently performed on clinical scanners using standard pulse sequences at multiple *b*-values (diffusion weightings) to distinguish signal from four discrete tissue micro-compartments (intracellular water, extracellular hindered water, freely diffusing water, and flowing fluid)^21–24^. Retrospective studies have shown the potential for RSI to make csPCa more visible and to improve radiologist accuracy^25,26^. RSI also lends itself to superior correction of DWI distortion (e.g., from rectal gas) that can interfere with MRI quality (and may be even more important for studies performed without contrast sequences as an image quality “safety net”)^21,27,28^. Beyond subjective interpretation, RSI yields a quantitative imaging biomarker, the RSI restriction score (RSIrs), that is superior to conventional apparent diffusion coefficient (ADC) for patient-level detection of csPCa^29,30^. The maximum RSIrs in the prostate can be determined automatically, without the need for a radiologist to first subjectively define a lesion of interest, and has been shown to perform similarly to expert PI-RADS interpretation for patient-level detection of csPCa^29^. These results have been replicated in a large study from the Quantitative Prostate Imaging Consortium, involving RSI data from 17 scanners and 7 imaging centers^30^. A prospective study also showed that non-radiologists were significantly more likely to correctly identify expert-defined csPCa on MRI when they were given RSIrs maps, compared to when they used conventional mpMRI^31,32^. As an objective biomarker, RSIrs could level the radiology playing field and facilitate more consistent interpretation of prostate MRI.

The objective of the Advanced Restriction imaging and reconstruction Technology for Prostate MRI (ART-Pro) study is to evaluate whether modern technologies can overcome two major barriers to widespread accurate prostate MRI: need for IV contrast and dependence on reader expertise. ART-Pro will be conducted in two stages. In ART-Pro-1, we will test whether IV contrast can be avoided in the setting of standardized, state-of-the-art image acquisition, with or without addition of RSIrs. This is a multisite study, and in contrast to most prior and ongoing studies, we seek to minimize variability of image quality by standardizing MRI acquisitions across all sites. We will also evaluate RSIrs as a stand-alone, quantitative, objective biomarker for detection of csPCa. In ART-Pro-2, we will measure the impact of RSIrs and IV contrast on the accuracy of *non-expert radiologists’* detection of csPCa. While ART-Pro-1 involves a select group of expert prostate radiologists at centers of excellence (to establish the gold standard), ART-Pro-2 is a pre-planned retrospective study of non-expert radiologists’ interpretations of ART-Pro-1 images.

## 2. Patients and Methods

### 2.1 Study Design

The ART-Pro study aims to evaluate prostate MRI techniques in two stages. ART-Pro-1 is a multisite, multinational, paired cohort trial evaluating whether IV contrast can be avoided in the setting of standardized, state-of-the-art image acquisition, with or without addition of RSIrs. RSIrs will also be evaluated as a stand-alone, quantitative, objective biomarker. ART-Pro-2 is a pre-planned retrospective, multisite, multinational study that leverages the (state-of-the-art, standardized) ART-Pro-1 dataset to study the impact of RSIrs and IV contrast on the accuracy of non-expert radiologists’ detection of csPCa. The trial is registered in the US National Library of Medicine Trial Registry (NCT number: NCT06579417). The expected trial timeline is three years to complete accrual with a six-month endpoint.

### 2.2 Objectives

The primary and secondary objectives are listed in Table 1.

**Table 1.** Study Objectives.

|  |
| --- |
| <u>Primary Objectives</u> |
| 1. Evaluate if bpMRI is non-inferior to mpMRI among expert radiologists (ART-Pro-1) and non-expert radiologists (ART-Pro-2) for detection of grade group (GG) $\geq 2$ csPCa |
| <u>Secondary Objectives</u> |
| 1. *Evaluate if RSIs plus axial $T_2$-weighted MRI is non-inferior to mpMRI among expert and non-expert radiologists for detection of GG $\geq 2$ csPCa |
| 2. *Compare quantitative RSIs (objective interpretation) to qualitative mpMRI (subjective radiologist interpretation) for detection of GG $\geq 2$ csPCa |
| 3. *Evaluate bpMRI, mpMRI, and RSIs for avoidance of unnecessary biopsies (i.e., any biopsy resulting in no cancer or only GG 1) |
| 4. Evaluate bpMRI, mpMRI, and RSIs for detection of GG $\geq 3$ cancer |
| 5. Assess performance of the above MRI techniques for detection of each: GG 1 (overdiagnosis), GG 2, GG 3, and GG 4-5 |
| 6. Evaluate the quality of scan images |
| a. Percentage of cases with diagnostic quality imaging per PI-QUAL |
| b. Percentage of cases with moderate or severe distortion |
| i. Percentage of cases where distortion correction is useful |
| 7. Evaluate csPCa detection by targeted vs. systematic biopsy cores |
| 8. Measure inter-reader reliability of bpMRI and mpMRI |
| 9. Evaluate accuracy of radiologists' and RSIs-based estimates of overall probability of csPCa on biopsy |
| 10. Assess influence of RSIs on bpMRI interpretation |

\* Key secondary objectives

### 2.3 Study Population

We will acquire subject data from 5 sites: University of California San Diego (UCSD), University of California San Francisco (UCSF), Massachusetts General Brigham Hospital (MGB), Weill Cornell Medical College (Cornell), and the University of Cambridge, UK (Cambridge). Patients referred for a multiparametric prostate MRI at any of the five participating sites, with a clinical suspicion of prostate cancer and that meet all the inclusion/exclusion criteria in Table 2, will be eligible for the trial. We will include a total of 500 patients in the trial (100 from each of the five sites).

**Table 2.**
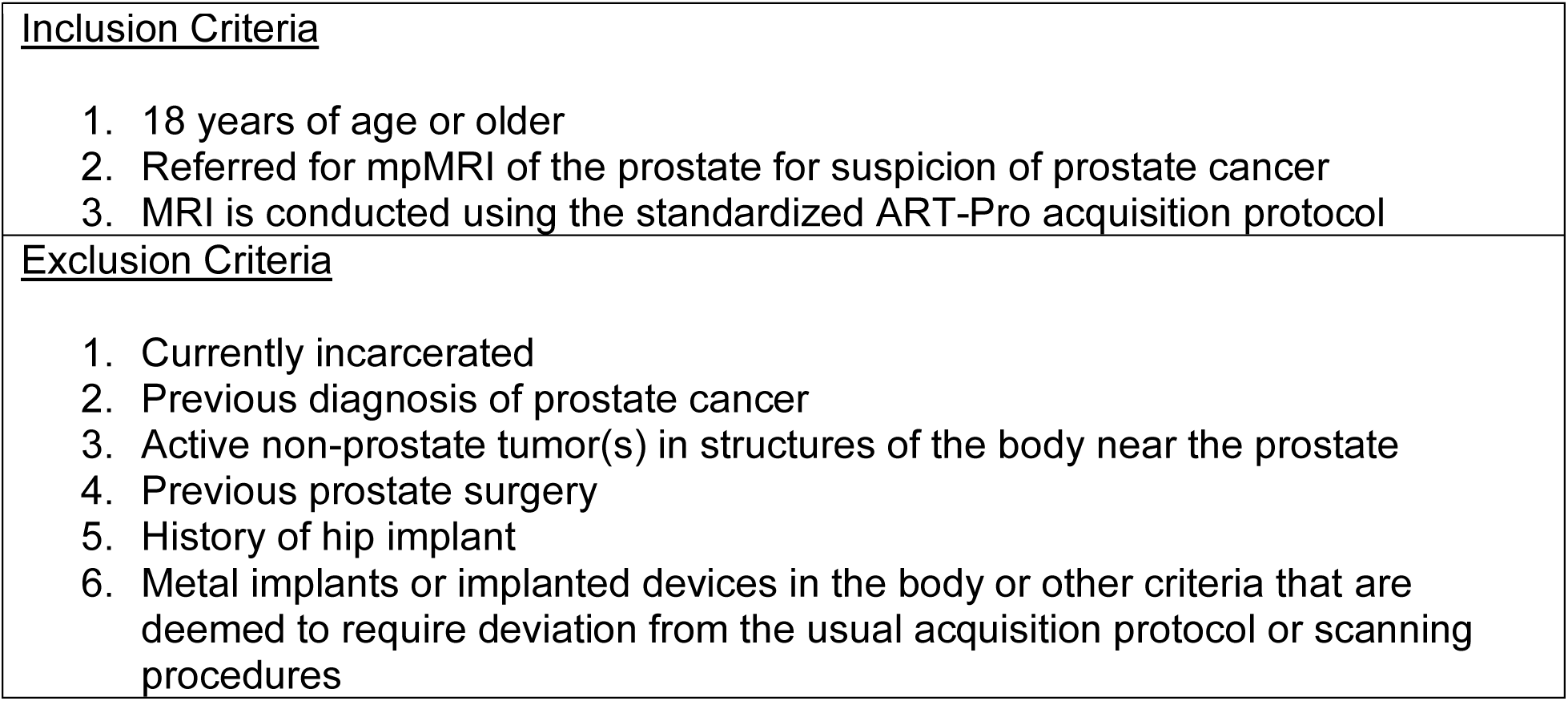
Eligibility Criteria.

|  |
| --- |
| <u>Inclusion Criteria</u> |
| 1. 18 years of age or older<br>2. Referred for mpMRI of the prostate for suspicion of prostate cancer<br>3. MRI is conducted using the standardized ART-Pro acquisition protocol |
| <u>Exclusion Criteria</u> |
| 1. Currently incarcerated<br>2. Previous diagnosis of prostate cancer<br>3. Active non-prostate tumor(s) in structures of the body near the prostate<br>4. Previous prostate surgery<br>5. History of hip implant<br>6. Metal implants or implanted devices in the body or other criteria that are deemed to require deviation from the usual acquisition protocol or scanning procedures |

### 2.4 Patient Recruitment

Patients are referred for a multiparametric prostate MRI for suspicion of prostate cancer per clinical routine. These patients are screened for study eligibility, and eligible patients are included in the study. The total scanner time for the ART-Pro study is comparable to routine clinical prostate MRI exams, and having two expert radiologists independently and collectively interpret the images would not be expected to adversely affect clinical care. Hence, at each of the four U.S. sites, local institutional review boards determined this study is HIPAA-compliant and could be conducted with a waiver of consent, based on minimal risk to patients. This recruitment approach also ensures patients included will be representative of the populations served by these institutions. At the University of Cambridge, approval has been secured by the local ethics committee to consent participants so that they can also give consent to sharing of data outside the UK.

### 2.5 Study Schema – ART-Pro-1

**Figure 1:**
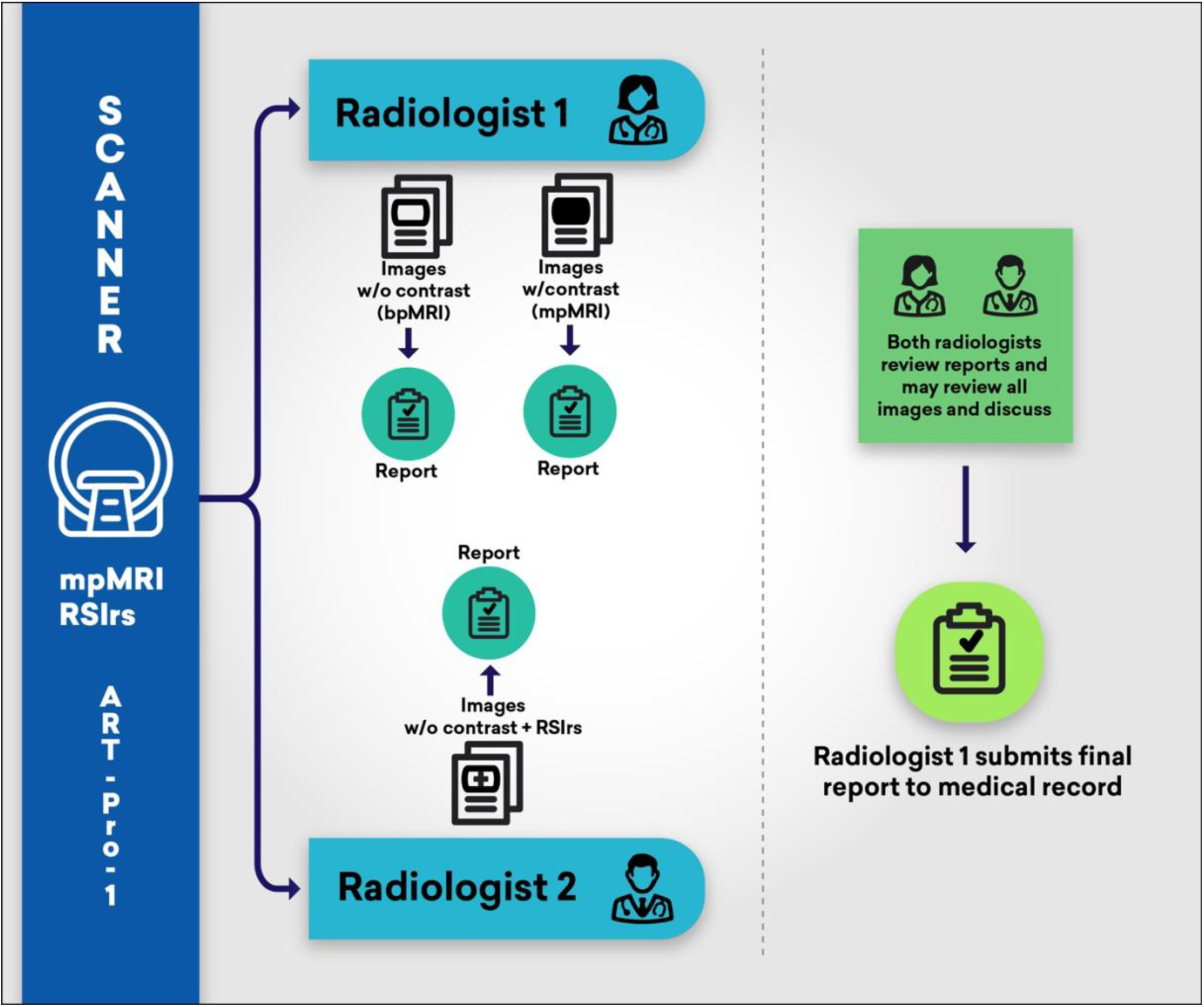
In ART-Pro-1, patients with suspected csPCa undergo MRI, with images interpreted independently by two expert radiologists. Radiologist 1 provides a research report using only bpMRI (no contrast) and then a second research report after reviewing DCE images (full mpMRI). Radiologist 2 provides a research report using bpMRI plus RSIrs. After each radiologist has submitted their research reports, they are given each other’s results. They may review all images and discuss the case, if useful. Radiologist 1 submits a final clinical report to the patient’s medical record to guide biopsy decisions.

### 2.6 MRI Acquisition and Technique

Eligible patients undergo mpMRI examination per clinical standard of care with additional images acquired for RSI. All examinations, in accordance with guidelines for standard mpMRI, include *T_2_*-weighted (T2W) sequences, diffusion-weighted imaging (DWI) sequences, and a dynamic contrast-enhanced (DCE) sequence. The additional images acquired for RSI add trivial time to the MRI examination and do not impose additional risk or increase billed charges to the patient. Acquisitions at 4 of the 5 sites will all be performed on the same 3T platform (SIGNA™ Premier XT, GE HealthCare, Waukesha, WI, USA) equipped with high-performance diffusion gradients, blanket surface coils (AIR™ Coils) and product deep learning-based denoising technology for *T_2_* and DWI (AIR™ Recon DL). Acquisitions at one of the 5 sites will be performed on a SIGNA™ Architect XT equipped with the same software and hardware except using slightly lower performance gradients.

The MRI acquisition protocol for ART-Pro was designed by consensus among the investigators, including physicists, engineers, and ten genitourinary radiologists from the five participating institutions, and with support from the scanner manufacturer. Development was informed by several empirical tests and votes for consensus (see supplemental material). Table 3 shows the imaging parameters in detail.

**Table 3.** MRI Parameters.

| | Acquisition time (min:sec) | $b$-values (s/mm<sup>2</sup>) [number of samples] | FOV<sup>1</sup>(m m) | Matrix | Slices | Slice thickness (mm) | TR<sup>2</sup> (ms) | TE<sup>3</sup> (ms) | TI<sup>4</sup>(m s) | Phases | Wash-in phases |
| --- | --- | --- | --- | --- | --- | --- | --- | --- | --- | --- | --- |
| **Localizer** |  |  |  |  |  |  |  |  |  |  |  |
| 3-Plane Localizer | 0:18 | N/A | 300x300 | 320x160 | N/A | 8 | Minimum | 80 | N/A | N/A | N/A |
| **$T_2$-weighted** |  |  |  |  |  |  |  |  |  |  |  |
| Axial $T_2$ | 2:07 | N/A | 160x160 | 360x224 | 32 | 3 | 3344 | 102 | N/A | N/A | N/A |
| Sagittal $T_2$ | 1:44 | N/A | 160x160 | 360x224 | 26 | 3 | 2717 | 102 | N/A | N/A | N/A |
| Coronal $T_2$ | 1:44 | N/A | 160x160 | 360x224 | 26 | 3 | 2717 | 102 | N/A | N/A | N/A |
| **$T_1$-weighted** |  |  |  |  |  |  |  |  |  |  |  |
| Axial $T_1$ | 0:47 | N/A | 200x200 | 256x256 | 1 | 3 | 5 | 2 | N/A | N/A | N/A |
| **Conventional DWI** |  |  |  |  |  |  |  |  |  |  |  |
| Axial reduced FOV (FOCUS<sup>5</sup>) | 1:53 | 50 [6], 1000 [18] (synth<sup>6</sup>1400 2000) | 160x88 | 100x50 | 32 | 3 | 4500 | Minimum | N/A | N/A | N/A |
| Axial extended FOV | 3:02 | 50 [6], 1000 [24] (synth<sup>6</sup>1400 2000) | 320x320 | 128x128 | 45 | 3 | 5088 | Minimum | N/A | N/A | N/A |
| **RSI DWI** |  |  |  |  |  |  |  |  |  |  |  |
| RSI with short TE | 3:25 | 0 [5], 100 [6], 800 [12], 1400 [12], 2500 [18] | 160x160 | 64x64 | 32 | 3 | 3800 | 80 | N/A | N/A | N/A |
| RSI with short TE and reverse phase-encoding polarity | 3:25 | 0 [5], 100 [6], 800 [12], 1400 [12], 2500 [18] | 160x160 | 64x64 | 32 | 3 | 3800 | 80 | N/A | N/A | N/A |
| RSI with long TE | 3:25 | 0 [5], 100 [6], 800 [12], 1400 [12], 2500 [18] | 160x160 | 64x64 | 32 | 3 | 3800 | 100 | N/A | N/A | N/A |
| **Dynamic contrast enhanced** |  |  |  |  |  |  |  |  |  |  |  |
| Axial DISCO<sup>7</sup> | 2:53 | N/A | 160x160 | 100x100 | 1 | 3 | 2.5 | Minimum | 23 | 38 | 32 |
| **Total scan time:** | 24:43 |  |  |  |  |  |  |  |  |  |  |

<sup>1</sup>Field of view
<sup>2</sup>Repetition time
<sup>3</sup>Echo time
<sup>4</sup>Inversion time
<sup>5</sup>Field-of-view Optimized and Constrained Undistorted Single-shot
<sup>6</sup>Synthetic *b*-value images, automatically computed from the acquired *b*-value data using built-in vendor software
<sup>7</sup>Differential Subsampling with Cartesian Ordering

Three RSI scans are included in the protocol for ART-Pro: two with a short echo time (TE) of 80 ms (the minimum achievable on the scanner hardware selected for the study) and one with a longer TE of 100 ms. The two short TE scans are acquired with opposite phase-encoding polarity (but are otherwise identical) to allow for effective correction of image distortions caused by inhomogeneities in the main magnetic field (*B_0_*)^27^. The RSIrs maps shown to radiologists during the study are the average of the RSIrs maps generated from the two opposite-polarity short TE scans. The long TE scan is not initially reviewed by any radiologist but is included to enable future investigations into the effects of *T_2_*-weighting on RSI signal properties and prostate cancer detection.

### 2.7 Innovation

Beyond standardization, the state-of-the-art MRI acquisition protocol used in ART-Pro incorporates several technological innovations. All *T_2_*-weighted and diffusion-weighted images in ART-Pro use high-density blanket surface coils and AI-based reconstruction to improve image quality and consistency. In addition to conventional DWI, ART-Pro includes multi-*b*-value DWI to permit calculation of RSIrs maps. The novel distortion correction method based on multi-*b*-value acquisition that is applied to RSIrs maps is used to improve cancer detection in cases where rectal gas leads to compression or stretching of prostate tissue on DWI^27^.

### 2.8 Image Distribution and Processing

Images from within each institution’s health IT network are transmitted from their scanner(s) to a processing/routing system created for ART-Pro. This system creates and processes two subsets of images, one for Reader 1 and one for Reader 2. The subset for Reader 1 includes the standard of care mpMRI sequences (DWI, T2W, and DCE). These images are fully identifiable with the patient ID and are no different than standard of care images when sent to PACS. The subset for Reader 2 includes bpMRI (without DCE) and RSIrs; these are de-identified and assigned an anonymized ID before being sent back to PACS. This de-identified worklist for Reader 2 ensures Reader 2 does not inadvertently view the full set of mpMRI images and/or interfere with Reader 1’s clinical worklist essential to clinical reporting. The processing system generates RSIrs maps using internal MATLAB code developed by the investigator team and described previously^22,23,29,30^. Briefly, the multi-direction (tensor), multi-*b*-value diffusion-weighted images are corrected for image distortions arising from *B_0_*-field inhomogeneity, gradient nonlinearity, and eddy currents. Background noise and receiver coil bias is then removed. The corrected data are fit to a multicompartment RSI model, and the signal from the slowest diffusion compartment is normalized by the median signal within the prostate on the *b*=0 s/mm^2^ images to generate RSIrs maps. RSIrs maps from both short TE scans are averaged together to generate the final RSIrs map that is distributed to the radiologists.

### 2.9 MRI Interpretation in ART-Pro-1

MRI exams are evaluated by two expert radiologists at the imaging center, with each radiologist interpreting approximately half the cases in the role of Reader 1 and half in the role of Reader 2 (either by random assignment or by which radiologist happens to be on clinical service the day the patient is scanned).

Reporting of MRI exams is done per PI-RADS v2.1 guidelines. Both readers also assess the quality of each modality reviewed (T2W, DWI, and DCE). Both readers know the clinical indication for the prostate MRI exam given by the ordering physician. Both also have access to the patient’s electronic medical record for age, race, ethnicity, family history, PSA level, prior biopsy information, etc. Both readers record whatever clinical information they reviewed by copying it into their research reports in our centralized REDCap database.

#### Reader 1

Reader 1 fills out a research report (Reader 1 Report), captured and stored in our centralized REDCap database, of the MRI findings. Reader 1 is first blinded to the DCE sequence and reports the MRI using only the biparametric (T2W and DWI) sequences. After reporting the bpMRI, Reader 1 is unblinded to the DCE sequence and re-reports the MRI using the full mpMRI (T2W, DWI, and DCE sequences). Reader 1 cannot go back and change their response for their bpMRI findings, so they ultimately provide two research reads: one without DCE and one with DCE. For both the bpMRI and mpMRI reports, Reader 1 provides an overall estimate of the probability of csPCa per lesion and per patient.

#### Reader 2

While blinded to the findings of Reader 1, Reader 2 completes a separate research report using bpMRI and RSIrs (Reader 2 Report), captured and stored in our centralized REDCap database. Lesions are assessed according to PI-RADS v2.1 with bpMRI; additionally, Reader 2 reports the maximum RSIrs for each lesion. Reader 2 also provides an overall estimate of the probability of csPCa per lesion and per patient.

As described above, RSIrs images in ART-Pro are acquired in two opposite phase encoding directions to permit multi-*b*-value correction of distortion due to *B_0_* field inhomogeneity. Both readers are asked to indicate whether the DWI is significantly distorted in each exam. If there is significant distortion in standard DWI, Reader 2 also reports whether the distortion is meaningfully reduced in distortion-corrected RSI.

#### Re-Evaluation

Once both Readers have completed their separate research reports, Reader 2’s report is delivered to Reader 1, the clinical radiologist of record. Upon review of Reader 2’s report and after any warranted discussion with Reader 2, Reader 1 has the opportunity to update their interpretation after considering Reader 2’s findings. Reader 1 fills out a second research report (Re-Evaluation Report), captured and stored in our centralized REDCap database, indicating if they would like to make any changes to their initial report. Reader 1 then submits a final clinical report to the patient’s electronic medical record.

#### Clinical Outcomes

Biopsy recommendations and other clinical decisions will be made by the patient’s medical team per clinical routine. We will review patient medical records to extract relevant outcomes, including whether a biopsy was recommended, whether a biopsy was performed (including technique), and the outcome of any biopsy: number of systematic cores and their locations; number of targeted cores and the corresponding target location; Gleason score and percentage of Gleason patterns per core; ductal or acinar type for any carcinoma; presence of cribriform pattern; presence of intraductal carcinoma; and presence of perineural invasion. If clinical interpretation includes notes of other poor prognostic pathology features, we will also record those^33^. If clinical genomic, pathomic, or other tests are performed on the tumor specimen (e.g., Decipher, Artera), we will record those results. If the patient undergoes radical prostatectomy, the final pathology will be recorded, including Gleason score, percentage of Gleason patterns, perineural invasion, tumor features (acinar, ductal, intraductal carcinoma, cribriform pattern), extraprostatic extension, seminal vesicle invasion, etc.

### 2.10 Study Schema for ART-Pro-2

**Figure 2:**
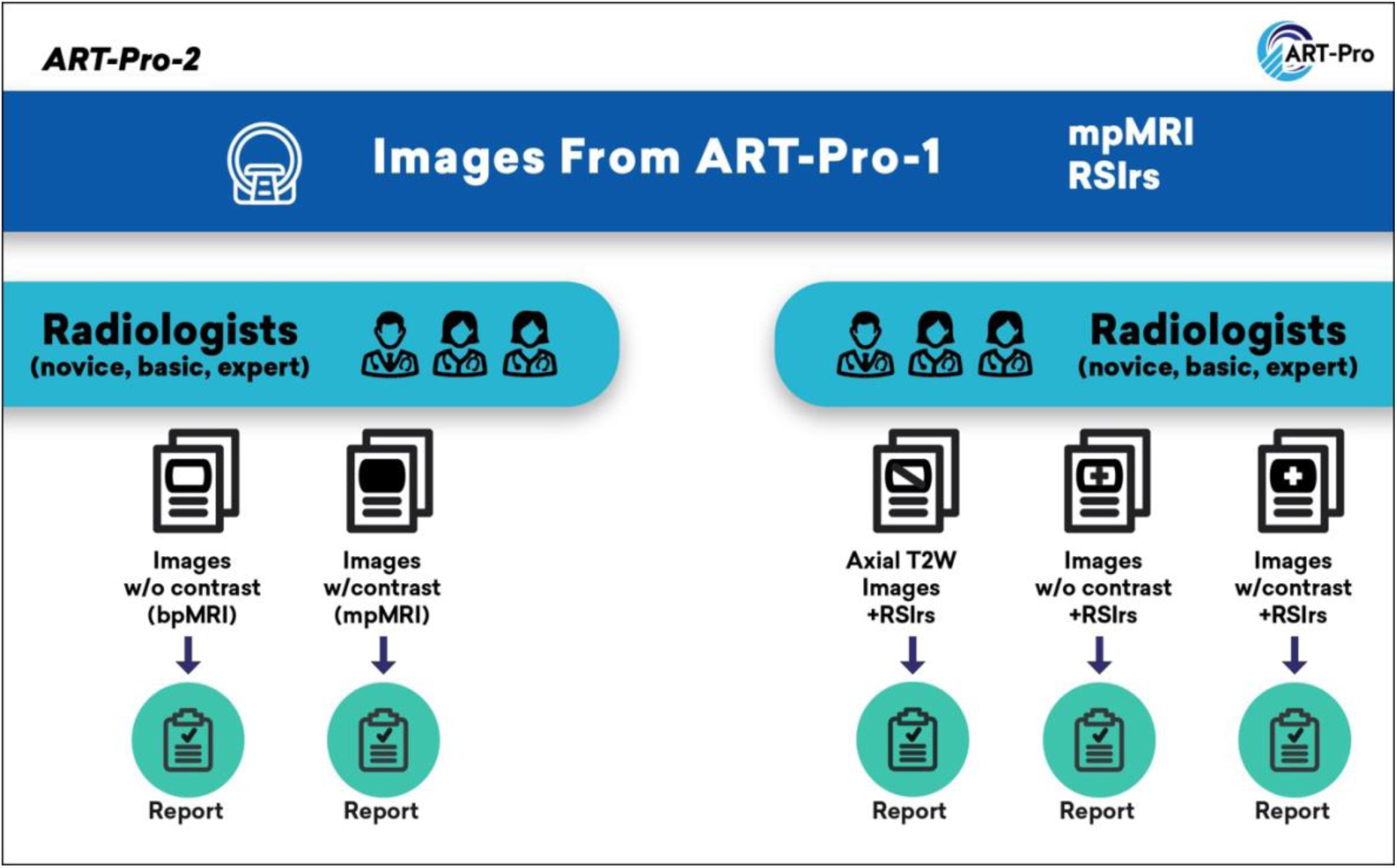
In ART-Pro-2, radiologists of different levels of experience (novice, basic, and expert according to ESUR/ESUI criteria) evaluate patient images from ART-Pro-1. For patient cases where they are assigned to mpMRI (left), the radiologists provide a report using only bpMRI (no contrast) and then a second report after reviewing DCE images (full mpMRI). For patient cases where they are assigned to RSIrs plus mpMRI (right), the radiologists provide three reports, in order: (1) based only on axial *T_2_*-weighted (T2W) images and RSIrs maps, (2) bpMRI plus RSIrs maps, and (3) full mpMRI plus RSIrs maps.

### 2.11 MRI Interpretation in ART-Pro-2

ART-Pro-2 will be conducted retrospectively and will not impact patient care. Radiologists will be categorized by level of experience for prostate MRI based on the joint European Society of Urogenital Radiologists (ESUR) and the EAU Section of Urological Imaging (ESUI) criteria: novice prostate radiologists defined as having read <400 cases, basic prostate radiologists defined as having read ≥400 and <1000 cases and expert prostate radiologists defined as having read ≥1000 cases^11^. The design of ART-Pro-2 in evaluating non-expert readers retrospectively allows for “locking” of reader experience level, whereas if evaluated over the ART-Pro-1 trial period, readers could change (by criteria) from being novice to basic and/or to expert. Radiologists will each be provided a list of 100 patient cases to review, 50 with mpMRI and 50 with RSIrs plus mpMRI. Case lists will be generated as random permutations of cases while requiring (1) that no radiologist be assigned cases from the institution where they work (to ensure they have not seen the cases before), (2) that each patient case is assigned to at least three radiologists as mpMRI (one from each of the experience levels), and (3) that each patient case is assigned to at least three (different) radiologists (one from each of the experience levels) as RSIrs plus mpMRI. Thus, each patient case will be reviewed by six radiologists (two from each of the experience levels). Image sets will be provided as sessions using the MIM Zero Footprint™ platform (MIM Software, Cleveland, OH) to facilitate individual worklists and presentation of image subsets.

#### mpMRI Patient Cases

For mpMRI patient cases, the radiologist will first be presented with bpMRI images and will complete a REDCap report with their findings according to PI-RADS v2.1. They will then proceed to review the DCE images for the case and will complete a second REDCap report according to PI-RADS v2.1. In each report, the radiologist will provide an estimate of the probability of csPCa per lesion and per patient.

#### RSIrs plus mpMRI Patient Cases

For RSIrs plus mpMRI patient cases, the radiologist will first be presented with only RSIrs maps and axial *T_2_*-weighted images. They will complete a REDCap report based only on images from these two series. As PI-RADS does not apply to RSIrs plus axial *T_2_*-weighted images, these reports will use the 5-point Likert scale used in the PROMIS trial: highly unlikely (1), unlikely (2), equivocal (3), likely (4), or highly likely (5). They will then proceed to review the full bpMRI image set (i.e., adding conventional DWI and ADC) and will complete a second REDCap report according to PI-RADS v2.1 guidelines. Finally, they will review DCE images and complete a third REDCap report for mpMRI according to PI-RADS v2.1. In each report, the radiologist will provide an estimate of the probability of csPCa per lesion and per patient.

## 3. Statistical Considerations

We expect bpMRI to be non-inferior to mpMRI among expert radiologists for detection of csPCa and avoidance of unnecessary biopsies. We expect expert radiologists will have superior performance to non-experts with bpMRI and with mpMRI. However, we hypothesize that adding RSIrs to bpMRI will facilitate objective MRI interpretation so that non-expert radiologists using RSIrs plus bpMRI will have non-inferior performance to experts using bpMRI or mpMRI. We expect RSIrs as a stand-alone quantitative biomarker will be non-inferior to qualitative mpMRI for discriminating patients with csPCa from those without csPCa.

### 3.1 Analyses for ART-Pro-1

#### Statistical analysis plan for primary and key secondary objectives

Exploratory data analysis will be conducted and visualization tools, including the scatterplot, boxplot and histogram, will be used to examine the data and potential missingness^34^. For all tests below, the significance level of 0.05 will be used in the statistical testing. All analysis will be conducted using software R 4.3.1.

For examining the non-inferiority of bpMRI vs. mpMRI for detection of csPCa, we will conduct one-sided non-inferiority tests of correlated proportions, to examine the sensitivity and specificity of the following paired readings: Reader 1 bpMRI vs. Reader 1 mpMRI and Reader 2 bpMRI vs. Reader 1 mpMRI. For both paired comparisons, the outcome of each reading for each subject is the binary result of positive or negative MRI. A McNemar’s test will be used to test the null hypothesis that the differences in the probabilities (% of second test minus % of the first test) of positive MRI (for testing on sensitivity) and negative MRI (for testing on specificity) from the two readings in the paired comparison are larger than the pre-determined non-inferiority margins^35^. P-values will be reported for all tests.

For examining the non-inferiority of RSIrs vs mpMRI, we will compare the areas under the receiver operating characteristic curve (AUCs) of using the RSIrs (continuous variable) to predict the binary outcome of csPCa vs. using the mpMRI (ordinal categorical variable; treat like continuous variable) to predict the outcome^36^. A non-inferiority test will be conducted to compare the two ROC curves associated with the two predictors at significance level 0.05.

#### Sample size calculations

All power calculations are conducted in software PASS 14.0.9 and R 4.3.1, unless otherwise indicated.

For testing the sensitivity, we expect the sensitivity to be 90% from the second test in each of the paired comparisons (Reader 1 mpMRI) and assume a margin of 8% in defining non-inferiority and an actual difference of 0%. We expect the percentage of probability of the first test giving a positive result while the second giving a negative result will be 5%. A sample size of 145 subjects achieves at least 80% power at significance level 0.05. We plan to enroll 500 patients and expect 30% will be diagnosed with csPCa, yielding 175 analyzable patients for evaluation of sensitivity. Given the design of the trial, missing data is expected to be low (3%). Therefore, with the proposed sample size, we will have ample power in the above statistical tests.

For testing the specificity, we expect the specificity to be 40% from the second test in each of the paired comparisons and assume a margin of 10% in defining non-inferiority^37^ and an actual difference of 0%. We expect the percentage of probability of the first test giving a negative result while the second giving a positive result will be 5%. Of 500 patients enrolled, 350 are expected to <u>not</u> be diagnosed with csPCa (either no biopsy due to low clinical/imaging risk or biopsy negative for csPCa), and missing data rate of 3%. A sample size of 340 analyzable patients achieves at least 80% power at significance level 0.05.

For testing the non-inferiority of the ROC curves associated with RSIrs and mpMRI, a preliminary study of 476 patients indicates the AUC of the model with RSIrs is 0.747 (95% CI: 0.7025-0.7884) while the AUC of the model with mpMRI is 0.767 (95% CI: 0.726-0.8085). With the proposed sample size, we will have at least 80% power in the non-inferiority test with a margin of 8%. The power analysis is conducted with R 4.3.1 and package pROC.

### 3.2 Analyses for ART-Pro-2

Statistical analyses for ART-Pro-2 will be analogous to those described for ART-Pro-1. In ART-Pro-2, each radiologist will give sequential reports on each assigned patient case: i.e., either [1] bpMRI and [2] mpMRI or [3] RSIrs plus *T_2_*-weighted axial, [4] RSIrs plus bpMRI, and [5] RSIrs plus mpMRI. We will compare each approach [1], [3]-[5] to mpMRI in analogous manner to in ART-Pro-1 using McNemar’s test and AUCs. These analyses will be performed within each stratum of ESUR radiologist expertise, testing for non-inferiority compared to mpMRI PI-RADS for ESUR-defined experts (essentially a validation of ART-Pro-1), radiologists with basic prostate MRI proficiency, and novices. In terms of statistical power, each of these ART-Pro-2 subgroups will have the same statistical power as the primary and key secondary analyses of ART-Pro-1.

Additionally, we will estimate the effect of radiologist expertise for each approach [1]-[5] in linear mixed effects models that utilize all data from ART-Pro-2. Models will take the form

csPCa status ∼ score + experience + (1|patient) + (1|radiologist)

where csPCa status is whether the patient was diagnosed with csPCa (binary), score is the PI-RADS or Likert score assigned, and experience is a categorical variable for ESUR/ESUI level of experience. Importance of radiologist experience within each approach [1]-[5] will be estimated by the parameter estimates for ESUR/ESUI basic proficiency and ESUR/ESUI novice levels, compared to a reference of ESUR/ESUI experts.

### 3.3 Secondary Analyses

We will repeat statistical analyses above using an alternate definition of GG ≥3 as true positives. GG 1 or benign will continue to be considered true negatives. We will also measure detection of cancers of each GG (GG 1, 2, 3, or 4-5). GG 1 diagnoses will be considered undesirable overdiagnoses. Rates of overdiagnosis will be compared by McNemar’s test, as above.

The percentage of cases with diagnostic quality imaging per PI-QUAL (as rated by Reader 1 in ART-Pro-1) will be measured for each site and for the study, overall. The percentage of cases with significant distortion of DWI will be measured for each site and for the study, overall. The percentage of such cases where distortion correction was deemed diagnostically helpful will be measured for each site and for the study, overall.

Inter-reader reliability of PI-QUAL scores and bpMRI PI-RADS scores will be summarized using Cohen’s kappa, comparing ART-Pro-1 Reader 1 to ART-Pro-1 Reader 2.

We will calculate the percentage of csPCa detected on systemic biopsy only, targeted biopsy only, or both.

Radiologists estimate the probability of csPCa on biopsy for each patient and record this in the REDCap forms. We have previously reported objective estimates of probability of csPCa for RSIrs maximum in the prostate^30^. For ART-Pro patients who undergo biopsy, we will construct ROC curves for radiologists’ estimates and for RSIrs estimates, with 95% confidence intervals via bootstrapping. We will compare discriminative performance for expert radiologists, non-expert radiologists, and RSIrs. We will also visualize calibration for each of these via qq plots^38^.

In ART-Pro-2, the overall performance of each approach [1]-[5] will also be compared by measuring the Akaike information criterion for each model.

## 4. Discussion

Prostate mpMRI improves the diagnostic pathway for prostate cancer by increasing detection of csPCa while also avoiding unnecessary biopsies. Major limitations of mpMRI are the need for IV contrast and its dependence on user expertise. The ART-Pro trial aims to test whether IV contrast can be avoided in the setting of standardized, state-of-the-art image acquisition, with or without addition of a quantitative imaging biomarker, RSIrs. Critically, ART-Pro will study whether novel technology can help non-expert radiologists achieve performance comparable to expert radiologists, even without use of IV contrast. Also, unlike other studies evaluating mpMRI and bpMRI for csPCa detection, ART-Pro employs fully standardized image acquisition protocols across multiple centers. Test characteristics of RSIrs will additionally be evaluated as a stand-alone, quantitative biomarker. The primary endpoint of ART-Pro is biopsy-confirmed csPCa. A key secondary endpoint is unnecessary biopsies (i.e., any biopsy resulting in no cancer or only GG 1).

ART-Pro is being conducted in two linked stages. ART-Pro-1 includes expert readers at five centers of excellence and will yield a carefully curated dataset under ideal conditions. ART-Pro-1 will answer several key questions: (1) Can IV contrast be avoided when expert radiologists are available? (2) Does use of RSIrs facilitate omission of IV contrast? (3) How does performance of RSIrs alone (as an objective quantitative biomarker) compare to expert bpMRI and mpMRI interpretation in a multi-center study? ART-Pro-2 will leverage the carefully curated dataset from ART-Pro-1 to investigate the impact of IV contrast and RSIrs maps, respectively, on the accuracy of non-expert radiologists. Since two of the drawbacks to prostate mpMRI (variable interpretation of results and false-positive findings) are exacerbated in non-expert radiologists, investigating these prostate MRI techniques in non-experts through ART-Pro-2 will be invaluable for determining utility of the techniques.

ART-Pro’s pragmatic design facilitates conduct of the study but also has some limitations. Biopsy decisions are made per clinical routine and are not prescribed by the study, so biopsy may not be performed for some patients despite a suspicious MRI. On the other hand, this reflects real-world practice. We expect conducting the study at centers of excellence will mitigate the risk of non-standard recommendations, and we have accounted for the possibility of patients declining a recommended biopsy in our power analysis. Clinical decisions for each patient are made based on the final recommendations of one radiologist (ART-Pro-1 Reader 1). It is obviously not possible to have the same patient undergo two separate decisions, and our design allows for both Reader 1 and Reader 2 to influence the biopsy decision and biopsy targets. An alternative approach would be a randomized trial where patients are only evaluated with either bpMRI, mpMRI, or RSIrs plus bpMRI. Such a randomized trial would suffer drawbacks of requiring many more patients (because of non-paired statistical comparisons) and a considerable consenting effort that would inevitably slow accrual and introduce biases because some patient populations are less able to participate in research studies (e.g., because of language barriers, complex wording of consent forms, burden of additional visits to complete consent, etc.). ART-Pro-2 has a retrospective design, meaning the ART-Pro-2 radiologists do not influence biopsy decisions. They may, for example, identify additional lesions that were not targeted on biopsy. Given that each patient can only have one biopsy recommendation and first biopsy procedure, we believe the most ethical and most accurate approach is to have two expert radiologists at a center of excellence influence biopsy recommendations that lead to the gold standard reference for the study’s primary outcome (presence of csPCa on biopsy).

ART-Pro has the potential to make several meaningful impacts on patient care. If bpMRI is proven non-inferior to mpMRI for detection of csPCa in the setting of standardized protocols and technology, patients could benefit from avoidance of discomfort and risks of IV contrast. This would also improve accessibility and decrease cost of pre-biopsy prostate MRI. Validation of RSIrs as a quantitative biomarker could improve objective interpretation and increase reproducibility of results for readers across a range of skill levels. If ART-Pro-2 demonstrates non-expert radiologists using only RSIrs and axial *T_2_*-weighted series have comparable accuracy to expert radiologists interpreting complete mpMRI, this would mean it may be possible to adopt a very short prostate MRI exam that could be widely implemented to significantly increase capacity for pre-biopsy MRI while ensuring reproducible and accurate results.

Additionally, the carefully curated dataset created in ART-Pro-1 will be useful for future prostate MRI studies. This dataset could be used, for example, to validate current and future AI-based prostate MRI tools and to measure their performance. Thus, ART-Pro will not only evaluate current protocols and technologies for prostate mpMRI but also has the potential to improve future prostate MRI research.

## 5. Supplement

### 5.1 Protocol design

For axial scans, participating institutions used a mixture of 16 cm and 20 cm FOV sizes. To select between these options, a volunteer was scanned using both FOV sizes (maintaining constant in-plane resolution in accordance with PI-RADS v2.1), and both sets of images were distributed to all participating radiologists for review. By majority vote, the 16 cm FOV was selected for axial imaging.

Axial *T_2_*-weighted images were also obtained from the same volunteer using two different acquisition strategies, cartesian fast spin echo (FSE) and radial “Periodically Rotated Overlapping ParallEL Lines with Enhanced Reconstruction” (PROPELLER) and compared by all participating radiologists. Cartesian FSE was selected by majority vote for inclusion in the protocol.

Imaging parameters for the other *T_2_*-weighted, *T_1_*-weighted, conventional DWI, and dynamic contrast-enhanced scans were determined via discussion among all participating radiologists and votes for consensus.

For RSI, resolution and scan coverage were selected for compliance with PI-RADS v2.1. *b*-values were determined from an estimation theory analysis described previously^39^.

## Data Availability

All data produced in the present study are available upon reasonable request to the authors.

